# Viral load monitoring practices and correlates of viral non-suppression among children and young adolescents living with HIV in level five hospitals in Kiambu county, Kenya

**DOI:** 10.1101/2024.06.03.24308381

**Authors:** Lilian N. Gachoka, Anne Njoroge

**Author notes:** **Corresponding author** Lilian N. Gachoka Email.

## Abstract

**Background:** HIV has been a major global public health issue among children and young adolescents living with HIV (CYALHIV), their viral suppression rates being lower compared to adults. Follow up through viral load monitoring may influence their health outcomes.

**Objective:** In a cross-sectional study we determined viral load monitoring practices and correlates of viral non-suppression of all CYALHIV in three level five hospitals in Kiambu county, Kenya

**Methods:** We abstracted data from electronic and paper medical records. Multivariable log binomial regression was used to estimate prevalence ratios (PR) and assess correlates of non-suppression. Adherence to viral load testing guidelines was assessed. Viral non-suppression was defined as a VL >1000c/ml.

**Results:** Of the 252 CYALHIV, the median age was 11 (IQR: 7-13) years. Fourteen had non-suppression at last assessment. Correlates of non-suppression included having previously had TB [aPR=4.25; 95% CI=1.41-12.8; p=0.01], ART side effects [aPR=3.01; 95% CI=1.37-6.62 p=0.006] and having received enhanced adherence counselling [aPR =5.32; 95% CI=2.00-14.15; p=0.001]. Being on Dolutegravir was significantly associated with a lower likelihood of non-suppression (aPR=0.35; CI:0.15-0.85: p = 0.021). Timing of baseline VL tests improved through the years though there were gaps in routine VL monitoring and follow-up on unsuppressed results.

**Conclusion:** At most current VL, 14% children were non-suppressed, higher than the 5% UNAIDS 2030 target. Special strategies on assessing and addressing corelates of non-suppression are essential for ART programs. Routine VL monitoring as per the guidelines was suboptimal despite increased access to VL testing, suggesting other barriers to VL monitoring.

## INTRODUCTION

In 2022, approximately 1.5 million children under 15 years were living with HIV globally, with the number of new infections in this age group projected to be 130,000, of which 45% were predicted to occur in Eastern and Southern Africa (1). In Kenya 7% of people living with HIV (PLHIV) are aged 15 years and below (2). It is imperative to target this age group with HIV treatment efforts aimed at reducing morbidity and mortality to safeguard their right to thrive.

Regionally, viral suppression rates amongst children and young adolescents living with HIV (CYALHIV) has been reported to be low(3) (4) (5) (6). In Kenya, prevalence of viral non-suppression among children was higher (51.8 %) than that observed in adults (28.4%)(7). Viral non-suppression is associated with increased risk of opportunistic infections and mortality(4) (5) Low level viremia has recently been shown to be associated with neurocognitive developmental delays (8) which could affect children’s learning capacity.

To achieve the 2030 UNAIDS 95-95-95 targets where 95% of PLHIV should know their HIV status, 95% PLHIV should be on ART and 95% of those on ART should have viral suppression, it is important to understand correlates of viral non-suppression to guide clinical practice and design of interventions.

Previously reported correlates of viral non-suppression among CYALHIV include being male, having had tuberculosis (TB), HIV status disclosure at an older age, experiencing ART adverse drug events, all of which have been associated with ART non-adherence (4)(9)(10). Some caregiver characteristics associated with viral non-suppression include one who isn’t the mother, caregiver substance abuse, poor socio economic status, inadequate social support (11) (12)(17).However, previous studies using routine programmatic data have been reported out of high HIV prevalence settings. It is important to explore whether these factors remain relevant in a low HIV prevalence setting.

WHO recommends routine HIV viral load (VL) monitoring as the preferred approach to ART monitoring. Routine VL monitoring could lead to earlier detection of treatment failure, with subsequent drug switches and optimized treatment regimens, with better treatment outcomes. It could also prompt drug resistance testing, which ultimately prevents empirical drug switches that could lead to drug resistance.

In Kenya, the 2018 guidelines on the use of ART for treating and preventing HIV recommended VL monitoring at treatment initiation and every six months thereafter. In case of virological non-suppression (VL > 1000 copies/ml) while on ART, enhanced adherence counselling (EAC) was recommended, followed by a repeat VL after three months. Treatment failure and resistance testing among CYALHIV ought to be confirmed before switching to second- or third-line regimens. However, the level of adherence to these guidelines in Kenya among CYALHIV is not well documented.

We set out to evaluate the prevalence and correlates of viral non-suppression and describe adherence to VL testing guidelines among CYALHIV in Kiambu County, Kenya, a region with low HIV prevalence and high access to VL testing.

## METHODS

### Study design

This was a cross-sectional study using data abstracted from electronic and paper medical records of CYALHIV in Kiambu, Kenya

### Study setting

Kiambu County is largely peri-urban and borders Nairobi, the capital city of Kenya. With an estimated population of 1,405 982 adults and 684,616 children below 15 years(14). It has a HIV prevalence of 2.1 % and an estimated 45917 persons living with HIV(15). It has 52 facilities offering comprehensive HIV care and treatment services serving both rural and urban populations, including the urban poor from neighboring slums in Nairobi.

We purposively selected three facilities (Kiambu County Referral, Thika and Gatundu Level Five Hospitals) because they have a high patient volume (3270, 4880 and 1665) active in care respectively and are located in distinct geographical semi urban setting with diverse populations. They are the main public referral hospitals in Kiambu county and have well established comprehensive care clinics (CCC). Most of the clinic services at these clinics are digitalized and use electronic medical records.

### Study population

All children and young adolescents living with HIV aged below 15 years who had been in care for at least six months and were active in care, marked by at least one clinic visit in the three months preceding data abstraction in October 2022 were included in the study. Children and young adolescents are typically reviewed by a clinician once every three months.

### Study Procedures

Variables of interest were structured into an electronic data capture tool (Kobo). Childs demographic characteristics, treatment /clinical factors and caregiver characteristics for caregivers who had HIV were abstracted from EMR system. Information for HIV negative caregivers was supplemented from paper records. Data abstraction was done between 3 rd October and 31 st October 2022. Research assistants were trained on the use of data collection instrument and on data abstraction from the records.

In these facilities, there is a sample referral system where whole blood samples are transported weekly to the national HIV reference lab for viral load testing.

### Definitions and correlates of viral non-suppression

Baseline viral suppression was defined as a viral load result within seven months of treatment initiation, while viral non-suppression was having ≥1000 copies per milliliter of plasma after being on ART for six months, (16). Undetectable VL was defined as having 0 copies/ml while 1-999 copies were classified as low-level viremia, both of which were considered viral suppression.

Covariates of viral non-suppression considered were the demographic characteristics including age, sex, education level, having siblings with HIV positive siblings and clinical/ treatment factors such as age at diagnosis, duration on ART, reported ART side effects, TB infection. Lack of adherence was defined as having received enhanced adherence counselling (EAC).

Caregiver characteristics evaluated included sex, relationship to the child and HIV status. For those who had HIV, age, education level, employment and viral suppression status were also included.

### Statistical analysis

We summarized categorical variables using counts and proportions while continuous variables were described using medians and interquartile ranges (IQR). Log binomial regression analysis was used to assess factors associated with viral non-suppression. Measures of association in form of prevalence ratios (PR) and a 95% confidence interval (CI) were obtained. Significant factors in bivariate analysis (p-value < 0.1) were then included in the multivariate analysis to obtain adjusted PRs. Analysis was done using STATA version 16 (Stata Corp, College Station, Texas, USA).

### Adherence to VL monitoring guidelines

Adherence to viral load testing was based on the national Kenyan 2012,2016 and 2018 guidelines for HIV treatment and prevention using the following four criteria’s: Baseline VL within 6 months of ART initiation, six monthly VL monitoring, enhanced adherence counselling and follow up VL monitoring in case of viral non-suppression and resistance testing after two consecutive non-suppressed VLs with EAC.

We also assessed the switch to Dolutegravir as an ongoing optimization drive was happening in Kenya between 2018 and 2022.

### Ethical considerations

The study was approved by Kenyatta National Hospital/University of Nairobi Ethics and Research Review Committee (P268/03/2022) and National Commission for Science, Technology and Innovation (NACOSTI/P/22/20290). Permission to carry out research was granted by County Department of Health Research Unit and the administration of participating hospitals. The Ethics committee gave waiver for individual patient consent as routinely collected data was anonymously abstracted from the medical records. Authors had no access to any information that could identify individual participants during or after data collection

## RESULTS

Of the 281 CAYLHIV active in care at the time of abstraction, we abstracted records from 252 who met the inclusion criteria (Figure 1). The median age was 11 (IQR: 7-13) years, and majority 198 (79%) diagnosed at the age of 0-4 years with 135 (54%) being male (Table 1). A majority 128 (51%) were from Thika.

**Figure 1.**
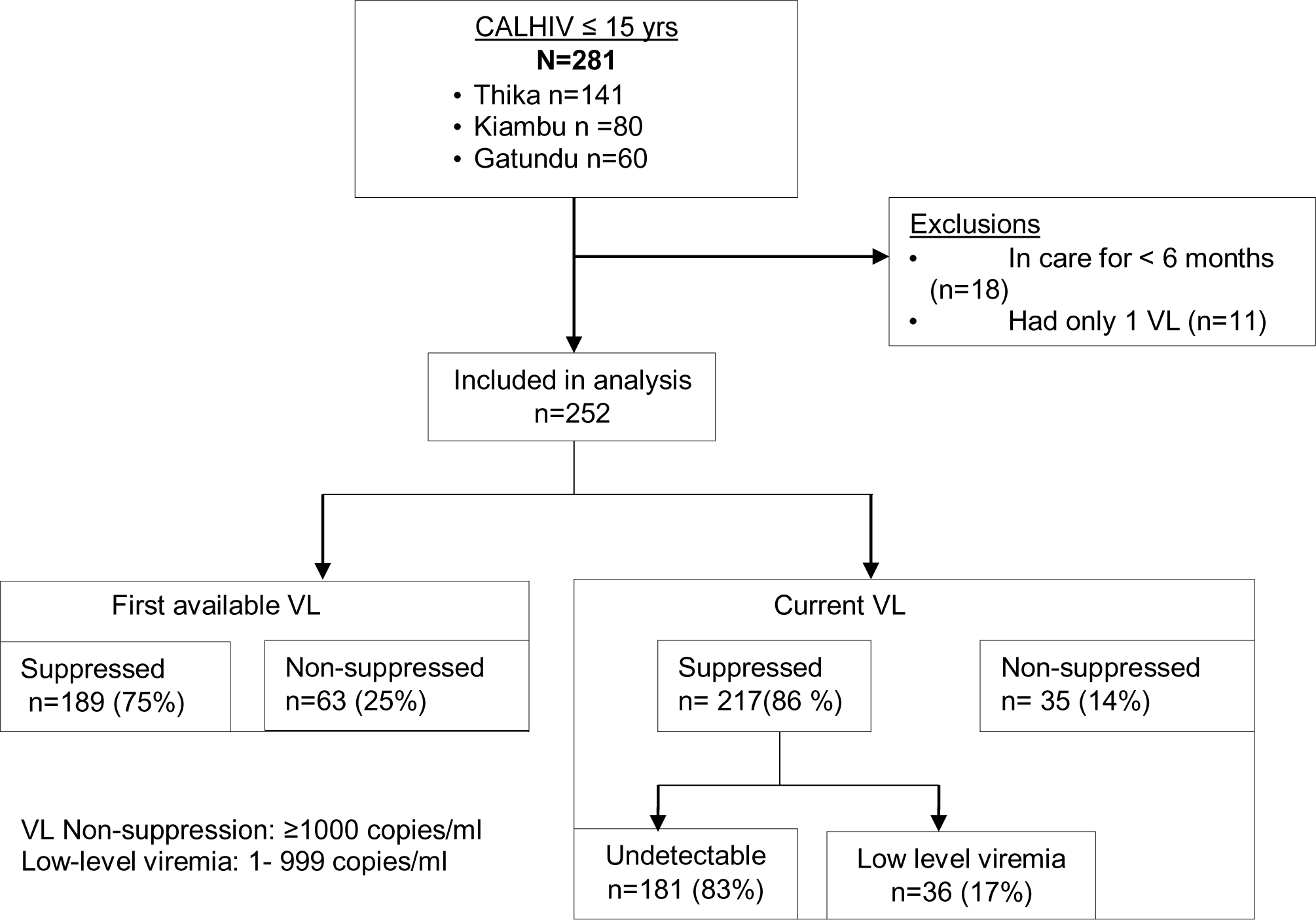
Prevalence of viral non-suppression.

**Table 1.** Characteristics associated with current viral non-suppression.

| Characteristic | n (%)<br>(N=252) | Bivariate Analysis |  | Multivariate analysis |  |
| --- | --- | --- | --- | --- | --- |
|  |  | Crude PR (95%CI) | p-value | Adjusted PR (95%CI) | p-value |
| **Current age (years)** |  |  |  |  |  |
| 0 - 4 | 30 (12%) | 1.87(0.86-4.04) | 0.114 | 1.02 (0.34-3.08) | 0.972 |
| 5- 9 | 70 (28%) | 1.03 (0.49-2.16) | 0.941 | 1.02 (0.42-2.51) | 0.961 |
| 10-14 | 152 (60%) | Ref |  | Ref |  |
| **Sex** |  |  |  |  |  |
| Male | 135 (54%) | 1.47(0.77-2.78) | 0.240 |  |  |
| **Education** |  |  |  |  |  |
| Nursery | 26 (10%) | 0.55(0.14-2.19) | 0.397 |  |  |
| Secondary | 31 (13%) | 1.15(0.48-2.79) | 0.749 |  |  |
| No school | 16 (6%) | 1.34(0.45-3.96) | 0.594 |  |  |
| Primary | 179(71%) | Ref |  |  |  |
| **Siblings with HIV** |  |  |  |  |  |
| Yes | 44 (18 %) | 0.87(0.38-1.98) | 0.735 |  |  |
| **Age at diagnosis (years)** |  |  |  |  |  |
| 0 - 4 | 198 (79%) | 1.01(0.91-1.14) | 0.800 |  |  |
| 5-9 | 48 (19%) |  |  |  |  |
| 10-14 | 6 (2%) | Ref |  |  |  |
| **Currently on DTG** |  |  |  |  |  |
| No | 14(6%) | Ref |  |  |  |
| Yes | 238(94%) | **0.24 (0.13-0.44)** | **<0.001** | **0.35(0.15-0.85)** | **0.021** |
| **ART side affects** |  |  |  |  |  |
| No | 230(91%) | Ref |  |  |  |
| Yes | 22 (9%) | **4.79 (2.72-8.42)** | **<0.001** | **3.01(1.37-6.62)** | **0.006** |
| **Tuberculosis** |  |  |  |  |  |
| No | 242(96%) | Ref |  |  |  |
| Yes | 10 (4%) | **3.12 (1.37-7.14)** | **0.007** | **4.25(1.41-12.8)** | **0.010** |
| **Enhanced adherence counselling** |  |  |  |  |  |
| No | 141(56%) |  |  |  |  |
| Yes | 111(44%) | **7.62 (3.05-18.9)** | **<0.001** | **5.32(2.00-14.2)** | **0.001** |
| **Duration on ART (years)** | 7((5-11) | 0.99 (0.99-1.00) | 0.078 | 0.99(0.99-1.01) | 0.715 |
| **Disclosure** | 166(66%) | 0.88 (0.46 1.72) | 0.72 |  |  |
| **Caregiver relationship\*** |  |  |  |  |  |
| Parent | 182(72.3%) | Ref |  |  |  |
| Grandparent | 20(7.9%) | 0.31(0.05-2.18) | 0.241 |  |  |
| Guardian | 6(2.5%) | 1 |  |  |  |
| Blood relative | 31(12%) | 0.61(0.20-1.87) | 0.385 |  |  |
| Sibling | 13(5.3%) | 0.97(0.26-3.61) | 0.958 |  |  |
| **Caregiver Sex\*** |  |  |  |  |  |
| Male | 34(14%) | 1.33(0.60-2.96) | 0.489 | 1.54(0.46-5.15) | 0.476 |
| **HIV positive caregiver (N=174)** |  |  |  |  |  |
| **Caregiver age (years)** |  |  |  |  |  |
| <40 years | 88 (51%) | Ref |  |  |  |
| >40 years | 86 (49%) | **0.44 (0.89-0.99)** | **0.014** | **0.44(0.19-0.98)** | **0.05** |
| **Caregiver education** |  |  |  |  |  |
| Primary | 92 (53%) | Ref |  |  |  |
| Secondary | 70 (40%) | 1.43 (0.69-2.93) | 0.336 |  |  |
| Tertiary | 4 (2%) | 1.92 (0.32-11.3) | 0.473 |  |  |
| Never attended school | 8 (5%) | 1.92 (0.52-7.11) | 0.331 |  |  |
| **Caregiver employment** |  |  |  |  |  |
| Employed | 22(13%) | Ref |  |  |  |
| Not employed | 152(87%) | 3.91 (0.56-27.3) | 0.17 |  |  |
| **Caregiver viral suppression** |  |  |  |  |  |
| Suppressed | 160 (92%) | Ref |  |  |  |
| Non-suppressed | 7(4%) | 1.76 (0.52-5.97) | 0.37 |  |  |
| Missing | 7 (4%) |  |  |  |  |
| *Only sex and relationship were available for caregivers who were not HIV positive. |  |  |  |  |  |

Majority, 218(87%) of the caregivers were females with 182 (72%) being the parent and 88 (51%) were less than 40 years old, with 174 (70%) of the caregivers also having HIV (Table 1)

### Prevalence of viral non-suppression

A quarter (63/252) had viral non-suppression at the first available VL, while the prevalence of non-suppression at the most current VL test, within a year of the date of data abstraction, was 14% (Figure 1)

### Correlates of viral non-suppression

As at October 2022, nearly all (94.4%) had been switched to Dolutegravir and had a lower likelihood of non-suppression. [adjusted PR [aPR]: 0.35 (CI:0.15-0.85:P = 0.021)]. Having reported ART side effects [aPR]:3.01; (CI:1.37-6.62: P = 0.006), history of TB [aPR]:4.25 (CI:1.41-12.8: P = 0.010), having received EAC [aPR]: 5.32 (CI: 2.00– 14.45; P= 0.001) were significantly associated with a higher risk of viral non-suppression. Children with older caregivers (>40 years) living with HIV were less likely to have non-suppression [aPR]: 0.44(CI:0.89-0.99; p value: 0.05) (Table 1)

### Adherence to VL testing guidelines

#### i. Baseline VL within 6 months of ART initiation

For the entire cohort, only 36 (14%) children and young adolescents had a baseline VL test within the recommended 6 (5-7 months) after treatment initiation which was the guideline since 2012. Before 2010, nearly all (80%) the baseline VL tests were done after 24 months. However, as time went on, more children were getting VL tests at baseline after ART initiation. After 2018, 61% of the children were tested within 6 months of ART initiation as recommended (Table 2)

**Table 2.** Time to baseline VL tests over the years.

| **Time to baseline** |  |  |  |
| --- | --- | --- | --- |
| **VL test** | **2010 -2015** | **2015 -2018** | **After 2018** |
| Upto 6 mths | 6(6%) | 11(14%) | 19(61%) |
| 6-12 mths | 5(5%) | 14(17%) | 5(16%) |
| 12-24 mths | 9(9%) | 20(24%) | 7(23%) |
| >24 mths | 119(80%) | 37(45%) | - |

#### ii. Six monthly VL monitoring

Restricting the analysis to viral load tests done after 2018 (n=169), routine monitoring for majority of the children was outside of the recommended follow up VL tests at every six months. The number of those who received VL testing every six months reduced over time, with non-compliance being 90% at the most recent VL compared to 69% at the earlier VL (figure 2).

**Figure 2.**
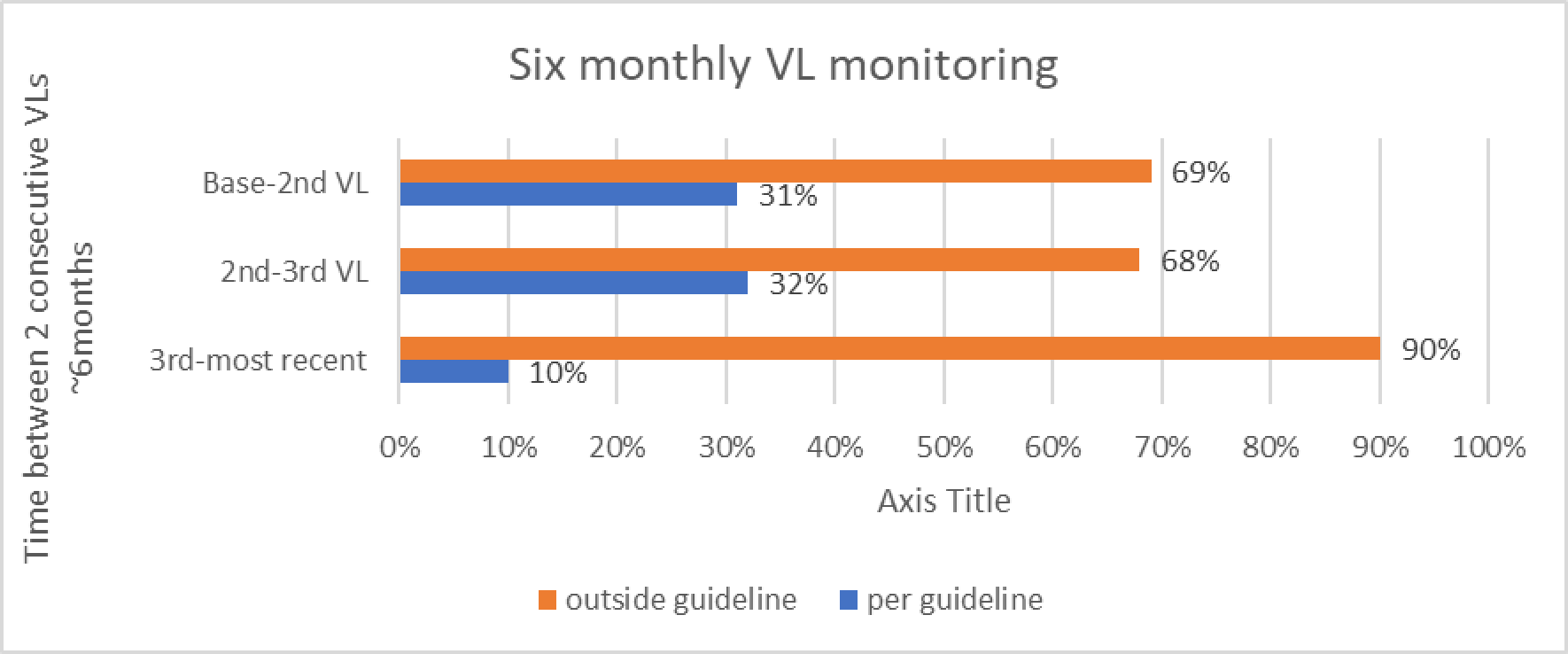
Routine VL monitoring within 6 months as per the 2018 guideline recommendation.

#### iii. Enhanced Adherence Counselling and follow up VL monitoring

Of the entire cohort, ninety-six CYALHIV had at least one non-suppressed viral load at one point in their medical history and the majority 77(80%) consequently received enhanced adherence counselling. Nevertheless, only 4(5%) children had a repeat VL test after the recommended three months of EAC (Figure 3)

**Figure 3.**
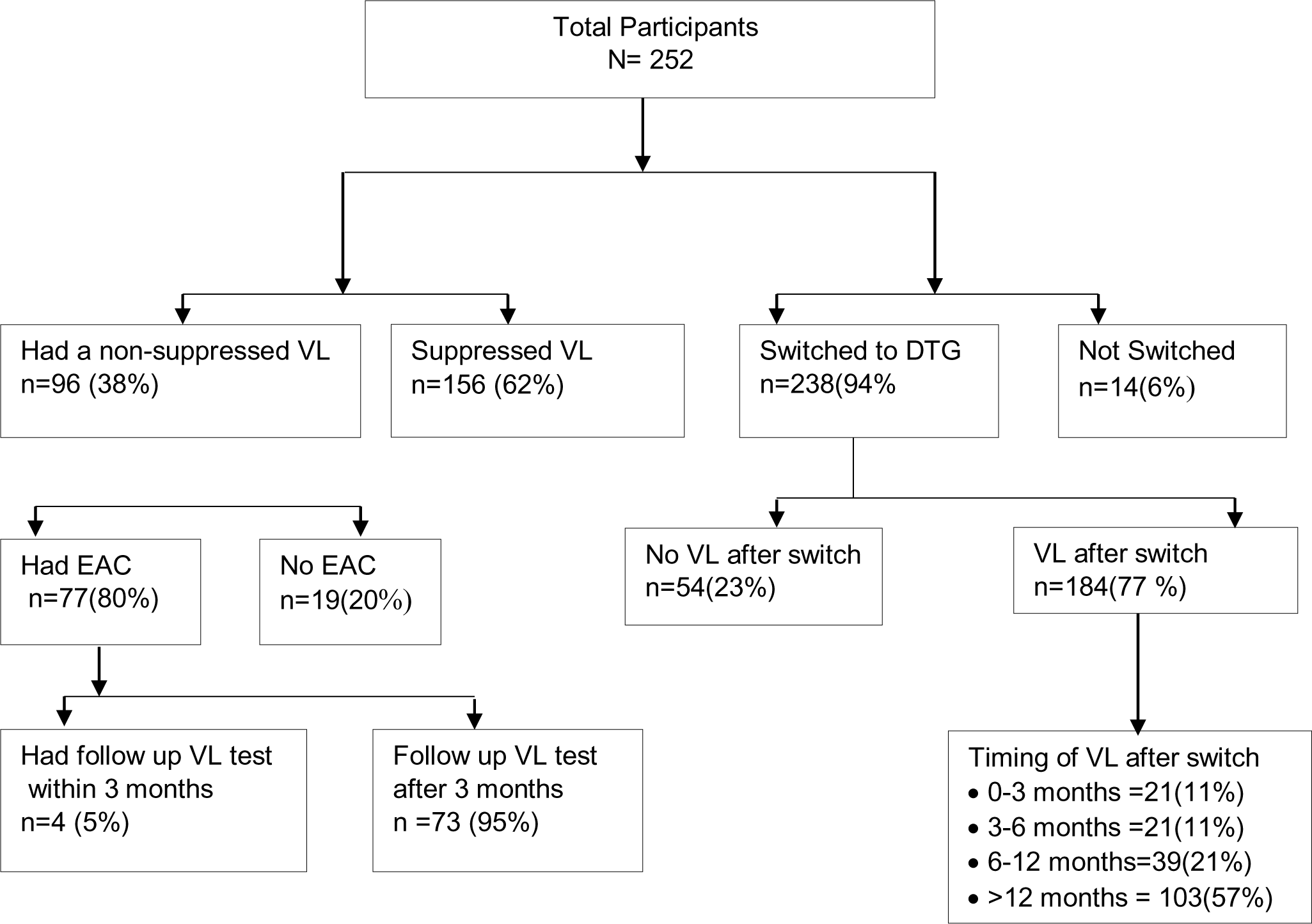
VL testing within DTG optimization.

#### iv. ART Drug Resistance testing

Looking at resistance testing for those children who had treatment failure, there were twelve CYALHIV who had been switched from a protease inhibitor (PI) to a DTG-based regimen due to viral non-suppression. Of these, only four had received drug resistance testing (DRT) as recommended in the 2018 guidelines. However, none of these DRT results were available in the EMR.

#### v. VL testing within DTG optimization

As at the time of abstraction, 238(94%) children had been switched to DTG. Of these (77%) had obtained a follow up VL after the switch. However, only a few 21(11%) were retested within the recommended time of three months (Figure 3).

## DISCUSSION

At most current VL, 14% of the CYALHIV had viral non-suppression. Correlates associated with higher likelihood of non-suppression included ART related side effects, had history of TB and having received EAC. Having an older (>40years) caregiver living with HIV was less likely to be associated with non-suppression. Viral load testing at treatment initiation improved over the years but routine monitoring was suboptimal and declined over time.

Viral suppression is crucial in reduction of negative outcomes among CYALHIV but the rates are worryingly low compared to adults regionally(4) (5) (6) (9). Similarly, in our study viral suppression was way below the UNAIDS 2030 targets that aims at achieving viral suppression in 95 percent of the HIV positive persons on treatment. However, prevalence of non-suppression from this study was lower in comparison to the 2018 Kenyan national estimates that was 51.7 % (7).This could be due to the continued roll out and training for health workers on the use of new treatment guidelines and regimen changes to optimization that has been attributed to better viral suppression. Additionally, among CYALHIV with a VL <1000c/ml 14% had low level viraemia that could increase the risk of virological failure (17) and neurocognitive developmental delays (8) consequently, affect their learning capacity. Hence the need for close follow for these children.

Consistent with other studies, we found that younger children (0-4 years) though not significant had a higher likelihood of viral non-suppression compared to older (10-14 years) CYALHIV (9) (18) (19) (20) (21).This could reflect the different challenges faced by younger children during treatment and care such as non-pleasant medication, dosing inaccuracies, treatment administration problems that results to insufficient drug levels, increased risk of drug resistance and eventual virologic failure (22) (23) (24) (25). In addition it can be due to the positive effects of disclosure among the older children (19). While the median age was 11years as significant proportion were diagnosed at an early age (0-4 years). This may mean that they had been perinatally infected.

Similar to previously reported correlates we found that CYALHIV who had history of TB and experienced ART related side effects had a higher likelihood of non-suppression(4) (26). This may be due to the increased pill burden, TB drugs interaction with protease inhibitors, while active TB can cause biologic effects of immune activation resulting in HIV viremia (27). ART adherence contributes to achievement of viral suppression thus EAC is recommended for PLHIV who have non-adherence issues or have detectable VL (11) (20) (28). Our findings showed about (44%) children had received EAC and were more likely to be non-suppressed. Similarly, low suppression rates were reported in Uganda following EAC (29).This result of non-suppression among children who had received EAC and had good adherence currently might indicate a possibility of drug resistance thus, need for timely drug resistance testing. Additionally, those children who had problems with ART adherence for long time could have been targeted for EAC sessions. It is essential that HIV programs should take suitable considerations on adherence counselling and support for PLHIV (30). Treatment optimization has been associated with better treatment outcomes since its more convenient, easy to administer, more tolerable palatable in children(31).This had been embraced from our findings as nearly all (94.4%) CYALHIV had been switched to DTG and had a lower risk of viral non-suppression

Studies have described the interaction between the child and their caregiver as a defining factor in the level of viral suppression for CYALHIV especially in resource limited situations(13).We found out that CAYLHIV with older HIV caregivers living with HIV had a lower risk of non-suppression. This may mean that older caregivers are more likely to remember the child’s ART and hence good treatment adherence. In addition, having a sibling/s living with HIV decreased the risk of non-suppression. This could possibly be due to treatment support they get from their siblings hence good adherence levels

WHO recommended routine VL monitoring has been reported as vital in early detection of ART failure, prevention of unnecessary switches thus improving treatment care outcomes (32)(33) (34)(35)(36)(37)(38)(39)(40)(41). The Kenya national ART guidelines from 2016 recommended VL monitoring at treatment initiation and every six months thereafter. It also recommended that children with a VL >1000 c/ml should receive EAC and a repeat VL test after 3 months of counselling. If the VL is unsuppressed, this confirms treatment failure and a switch to second line treatment while for children on a protease inhibitor (PI) based regimen, drug resistant testing is recommended (3).

The trend of time to baseline VL testing indicates improvement over time, during which VL testing became more accessible. However, only 61% of the children were tested within 6 months of treatment initiation as recommended on or after 2018, which was lower than expected. Routine VL monitoring was also suboptimal with time and health care providers adhering to the guidelines progressively reduced. This indicates other barriers to VL testing besides access, need to be explored. Some posited in literature include patients and staffs unawareness of benefits of VL testing, inadequate health care workers training, shortage of staffs, longer turnaround time of results and weak testing systems(32) (42) (43) (44) (45).

Several factors have been known to hinder ART adherence among CYALHIV. Support provided during the EAC sessions has previously been linked to improved treatment adherence resulting in higher viral suppression rates (46) (47) while averting occurrence of drug resistance (48). This was seen in this study as majority (80%) CYALHIV who had non-suppressed VL received EAC. However, the effectiveness of this EAC may not be clearly substantiated, since most of the children did not get a repeat VL after the EAC sessions, as per guidelines.

Gaps in the VL cascade of care such as delayed follow up VL, delayed or lack of regimen switch in virological failure has been reported as a hinderance to ART treatment success (49) (50). In our study, only 5% CYALHIV had a follow up test to confirm treatment failure. This was similarly reported in Lesotho, only 25% of patients with a first unsuppressed VL were managed correctly according to WHO guidelines. This indicates that in real life VL monitoring may be inadequate as some of unsuppressed test results do not result in any action or follow up(51). Given that VL testing is not cheap, it is imperative that proper follow-up of unsuppressed viral load results be done to ensure resources are well utilized, in addition to prevention of negative health outcomes for these patients.

Guidelines provide for drug resistance testing, a vital component in management of treatment failure (52) and subsequently prevents pragmatic drug switches that could lead to drug resistance. Studies have reported that upto 60-90% children who don’t attain viral suppression tend to have drug resistance (53) (54), especially mutations to PI drugs which may hinder success of HIV care interventions in children. In this study amongst the CYALHIV who had been switched from PI to DTG based regimen as a result of non-suppression, only 33% had received drug resistance. This indicates there is need for health care providers sensitization and clinical decision support in the care of children who might have ART failure.

Several limitations in our study include its cross-sectional nature and that we were using abstracted data, limiting our ability to enquire deeper into some of the findings as well as probable missing VL results, our key strength is that our study sites contained ∼85% of CAYLHIV in Kiambu county and therefore our results are generalizable to public facilities offering HIV care in low-prevalence HIV settings such as ours.

## Conclusion

At the most recent VL, 14% children were non-suppressed, higher than the 5% UNAIDS 2030 target. Special strategies on assessing and addressing corelates of non-suppression are essential for ART programs. Routine VL monitoring as per the guidelines was suboptimal despite increased access to VL testing, suggesting other barriers to VL monitoring, which need to be further explored, if we are to achieve epidemic control by 2030.

## Data Availability

N/A

## Acknowledgements

We acknowledge the study hospitals and Kiambu county research department for allowing us to collect data.

## Author contributions

L.N developed the study materials. L.N and A.N conceptualized and did data analysis. L.N wrote the first draft of the manuscript. A.N provided feedback and approved the final submitted manuscript.

## Abbreviations

ART, Antiretroviral therapy; CCC, Comprehensive care clinics; CDC, Centre for disease control and prevention; CYALHIV, Children and young adolescents living with HIV; DTG, Dolutegravir; DRT, Drug resistance testing; EID, Early infant diagnosis; EAC, Enhanced adherence counselling; HIV - Human immune deficiency virus; KENPHIA, Kenya Population-based HIV Impact Assessment; PLHIV, Persons living with HIV; UNAIDS, United Nations program on HIV/AIDS; UoN, University of Nairobi; VL, Viral load; WHO,World Health organization

